# High Prevalence of the Cardiovascular-Kidney-Metabolic Syndrome Among US Adults From 1999-2020 - An analysis of the NHANES survey

**DOI:** 10.1101/2024.03.04.24303751

**Authors:** Zhejia Tian, Samira Soltani, Johann Bauersachs, Kai Schmidt-Ott, Anette Melk, Bernhard MW Schmidt

## Abstract

**Background:** The cardiovascular-kidney-metabolic (CKM) syndrome is a newly defined chronic health condition from American Heart Association. We assessed the prevalence of CKM syndrome stages 0-2, which have not yet progressed to cardiovascular disease (stage 3-4) with trends analysis over the past two decades.

**Methods:** We used cross-sectional data provided by National Health and Nutrition Examination Survey. including non-pregnant participants aged 18 or older between 1999 and 2020. Weighted prevalence was analyzed over the course of the past 20 years and by population subgroup (including age, sex, and race/ethnicity).

**Results:** A total of 32848 US adults were included in our study (weighted mean age, 47.3 years; women, 51.3%). 7.9% of US adults were at stage 0 without any CKM risk factors, with 64% of this subgroup being female. 18.3% of US adults were classified as stage 1 with issues related to excess or dysfunctional adiposity without other metabolic risk factors or chronic kidney disease (CKD). More than half of the US adults (56.5%) exhibited either metabolic risk factors, CKD, or both (stage 2). Between 1999 and 2020, the CKM features increased with decreasing prevalence of stage 0 (P for trends =0.0018), not only in females but also in males.

**Conclusions:** Our findings illustrate an exceptionally high and increasing prevalence of CKM syndrome among US adults. This emphasizes the importance of comprehensive preventive strategies targeting the life style of large parts of the population. Moreover, further risk assessment should be implemented into stage 2 cohort to define patients with exceptional cardiovascular risk.

**Clinical Perspective:** *What is new?:*

- CKM syndrome is a common chronic health condition in the general population. However, the prevalence of different CKM stages using real-world data has not been reported within the general population or its subgroups.
- The prevalence of CKM syndrome was increasing over the past two decades. The majority of US adults were classified as stage 2.
- A specific population remained undefined according to the current detailed definition of each CKM syndrome stage.

*What are the clinical implications?:*

- The high and increasing prevalence of CKM syndrome necessitates more precise preventive strategies, tailored to different target groups with consideration of age-, sex-, and gender-disparities.
- Given that approximately half of the study population fell into stage 2 with a wide spectrum of risk factors, it is imperative to identify patients with exceptionally high risk through additional risk assessments. This approach would facilitate the implementation of intensified treatment measures aimed at preventing the progression to cardiovascular disease (CKM syndrome stages 3-4).

## Introduction

The cardiovascular-kidney-metabolic (CKM) syndrome is a recently defined chronic health condition characterized as a systemic disorder that signifies intricate interactions among metabolic risk factors, chronic kidney disease (CKD), and the cardiovascular system. It transcends the simple sum of its components, leading to multiorgan dysfunction and increased adverse cardiovascular outcomes ^1,2^. Four stages of CKM syndrome were outlined: stage 0, characterized by the absence of CKM risk factors; stage 1, marked by excess or dysfunctional adiposity; stage 2, involving metabolic risk factors and/or CKD; and stages 3-4, defined by subclinical (stage 3) or clinical (stage 4) CVD alongside CKM risk factors ^1,2^. Based on the latest heart disease and stroke statistics update, it is indicated that one in three adults in the United States has three or more risk factors contributing to the deterioration of CKM health. The overall prevalence of cardiovascular disease, including hypertension, coronary heart disease, heart failure, and stroke, stands at 48.6% among adults aged 20 and older ^3^. This highlights a substantial burden of CKM syndrome within the general population, underscoring the imperative for extensive healthcare initiatives.

Despite the significant challenges in managing poor CKM health, a comprehensive classification and risk assessment of CKM syndrome also provides us with opportunities to detect early stages for preventive measures and implement effective interventions, thereby decelerating progression. This is particularly important given the recent expansion of therapeutic innovations, such sodium-glucose co-transporter 2 inhibitors and glucagon-like peptide 1 receptor agonists ^4–8^. Additionally, optimizing CVD health necessitates the integration of social determinants, behavioral interventions, early-life prevention, multidisciplinary care, and ensuring affordable access to pharmacotherapy ^9–11^.

Extensive research and analyses were conducted to examine the impact of metabolic risks, CKD, and cardiovascular disease, along with their intersections ^3,12–16^. Despite these efforts, the prevalence of CKM syndrome, categorized by specific stages, remains undefined at this time. Using data from the National Health and Nutrition Examination Survey (NHANES), this analysis was intended to delineate the prevalence and distribution of CKM syndrome with a specific focus on stage 0-2 among US adults, aiming to identify subpopulations at risk for progression to stage 3-4 and to explore the trends of the prevalence over the last 20 years.

## Methods

### Data Source and Study Population

The NHANES has been conducted continuously in 2-year cycles since 1999. It employs a cross-sectional survey design to assess the health and nutritional status of adults and children in the United States, utilizing a complex, multistage, probability sampling design. The survey is representative for the non-institutionalized, civil population of the United States. Written informed content was obtained from all survey participants, and the study procedures receive approval from the National Center for Health Statistics Research Ethics Review Board. The Hannover Medical School Institutional Review Board exempted the present study because it did not constitute human subject research. We followed the Strengthening the Reporting of Observational Studies in Epidemiology (STROBE) reporting guideline throughout our study ^17^.

We included non-pregnant participants aged 18 or older from 10 NHANES cycles starting with the 1999-2000 cycle until 2017-March 2020 cycle in our analysis. All participants should possess sufficient information to determine cardiovascular disease based on self-report. All survey cycles were applied to evaluate the trends in the prevalence of CKM Stage 0-2 and were combined to analyze the overall prevalence.

### Data Collection

Demographic information was collected through household questionnaires. Race and ethnicity were not consistently reported in NHANES: non-Hispanic Asian participants were not classified and oversampled until 2011. In our analysis, we categorized self-reported Mexican American and other Hispanic individuals as Hispanic race and ethnicity. Data of Body mass index, waist circumference and blood pressure were available among participants who underwent clinical examination. Standardized blood pressure measurements were performed: three consecutive measurements were taken at one-minute intervals, and the average of the last two measurements was applied to our analysis ^18^. Hypertension was defined by either elevated blood pressure according to 2017 AHA guideline ^19^ or the use of antihypertensive medication.

Chronic kidney disease (CKD) is classified by estimated glomerular filtration rate (eGFR) and albuminuria according to KDIGO 2012 Guidelines as outlined in S1 Table ^20^. Urinary albumin to creatinine ratio (UACR) was extracted, if available, directly from medical examination data or was calculated from urinary albumin and urinary creatinine. We calculated eGFR using the 2021 race– and ethnicity-free Chronic Kidney Disease Epidemiology Collaboration creatinine equation ^21^.

For lipid profiles, we considered medication use based on self-report and laboratory examination of serum triglyceride, HDL-cholesterol and LDL-cholesterol. 2018 AHA guideline recommendations were applied to define the normal lipid condition ^22^.

We evaluated diabetes conditions using self-reported information on glucose-lowering therapy, medical examination of glycated hemoglobin and fasting serum glucose. The presence of diabetes or prediabetes was defined based on diagnostic tests outlined in the 2023 ADA guidelines ^23^

### Statistical Analysis

We first applied a selection strategy consistent with the definitions of CKM syndrome stage 0-2^1^ (S2 Table) to identify individuals meeting the criteria for stage 0-2 in each survey cycle and in overall combined cycles. We then accounted for the complex survey design factors for NHANES, such as sample weights, clustering, and stratification, as specified in the National Center for Health Statistic analytic guideline^24^. In all analysis, morning fasting subsample weights and Taylor series linearization method were applied to estimate prevalence and standard errors representative of the civilian, noninstitutionalized US population ^25,26^. Each prevalence estimate was reported with corresponding 95% confidence interval (95% CI). For trends analysis across cycles, survey-weighted logistic regression was employed, with survey cycle as a continuous variable. 2-sided *P* < 0.05 was considered statistically significant.

Sensitivity analyses were conducted to examine the influence of non-response on our main analysis. We first adjusted sample weights with the adjustment cell method ^27^. Moreover, multivariate multiple imputation by chained equations with 5 imputations was performed to address missing data^28^. All Analyses were conducted using R version 4.3.2.

## Results

### Study Population Characteristics

Over these 10 survey cycles, 32848 participants aged 18 or older were included in our final study, representing 215480397 noninstitutionalized US inhabitants (Table 1). Of all participants, mean (SE) age was 47.3 (0.2), 51.3% were female and 48.7% were male. Participants between 18 and 24 years old constituted 9.2% of the total, with 37.1% aged between 25 and 44, 35.6% between 45 and 64. 18.1% were aged 65 or older. More than 60% were Non-Hispanic White. Of the total, 13.8% identified as Hispanic, 11.1% as Non-Hispanic Black, 2.6% as Non-Hispanic Asian, and 4.5% reported as other race or ethnicity.

**Table 1.**
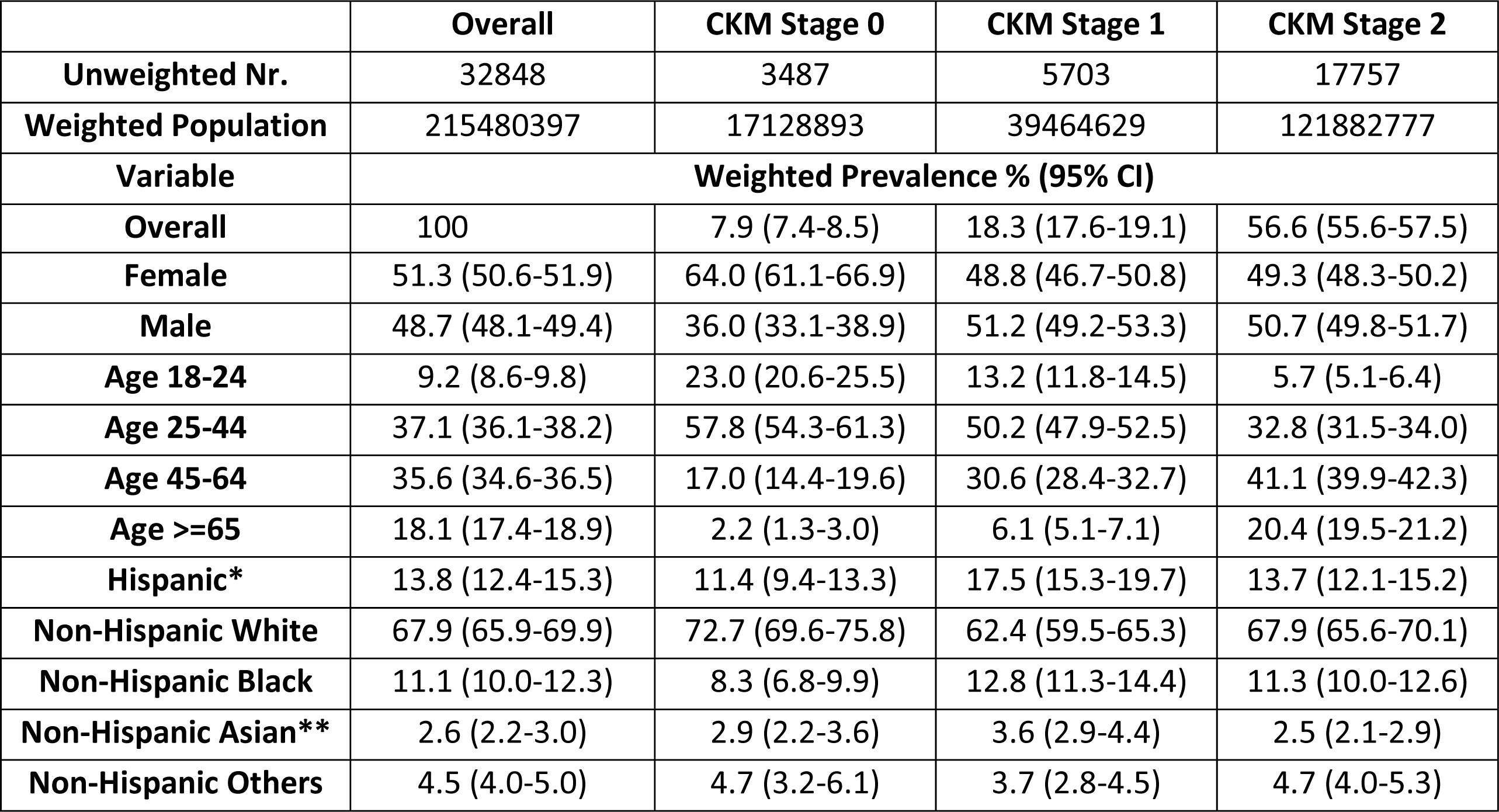
Weighted Population Characteristics Overall and by CKM Syndrome Stages in US Adults. Abbreviation: CKM, cardiovascular-kidney-metabolic syndrome. * Hispanic represents Mexican American and other Hispanic individuals. ** non-Hispanic Asian participants were classified and oversampled since 2011.

|  | Overall | CKM Stage 0 | CKM Stage 1 | CKM Stage 2 |
| --- | --- | --- | --- | --- |
| **Unweighted Nr.** | 32848 | 3487 | 5703 | 17757 |
| **Weighted Population** | 215480397 | 17128893 | 39464629 | 121882777 |
| **Variable** | **Weighted Prevalence % (95% CI)** |  |  |  |
| **Overall** | 100 | 7.9 (7.4-8.5) | 18.3 (17.6-19.1) | 56.6 (55.6-57.5) |
| **Female** | 51.3 (50.6-51.9) | 64.0 (61.1-66.9) | 48.8 (46.7-50.8) | 49.3 (48.3-50.2) |
| **Male** | 48.7 (48.1-49.4) | 36.0 (33.1-38.9) | 51.2 (49.2-53.3) | 50.7 (49.8-51.7) |
| **Age 18-24** | 9.2 (8.6-9.8) | 23.0 (20.6-25.5) | 13.2 (11.8-14.5) | 5.7 (5.1-6.4) |
| **Age 25-44** | 37.1 (36.1-38.2) | 57.8 (54.3-61.3) | 50.2 (47.9-52.5) | 32.8 (31.5-34.0) |
| **Age 45-64** | 35.6 (34.6-36.5) | 17.0 (14.4-19.6) | 30.6 (28.4-32.7) | 41.1 (39.9-42.3) |
| **Age >=65** | 18.1 (17.4-18.9) | 2.2 (1.3-3.0) | 6.1 (5.1-7.1) | 20.4 (19.5-21.2) |
| **Hispanic\*** | 13.8 (12.4-15.3) | 11.4 (9.4-13.3) | 17.5 (15.3-19.7) | 13.7 (12.1-15.2) |
| **Non-Hispanic White** | 67.9 (65.9-69.9) | 72.7 (69.6-75.8) | 62.4 (59.5-65.3) | 67.9 (65.6-70.1) |
| **Non-Hispanic Black** | 11.1 (10.0-12.3) | 8.3 (6.8-9.9) | 12.8 (11.3-14.4) | 11.3 (10.0-12.6) |
| **Non-Hispanic Asian\*\*** | 2.6 (2.2-3.0) | 2.9 (2.2-3.6) | 3.6 (2.9-4.4) | 2.5 (2.1-2.9) |
| **Non-Hispanic Others** | 4.5 (4.0-5.0) | 4.7 (3.2-6.1) | 3.7 (2.8-4.5) | 4.7 (4.0-5.3) |

### Overall Prevalence of CKM Syndrome Stages

The prevalence of different CKM syndrome stages with different population characteristics is summarized in Table 1. A total of 82.3% noninstitutionalized US adults were our target population, who would benefit from preventive cardiovascular assessments. Only 7.9% of the US adults showed no CKM risk factors and fulfilled the criteria for CKM syndrome stage 0, with the majority being female, comprising 64% of this subgroup. A higher percentage of younger participants were categorized in this group, with 74.9% being under the age of 45 years. 18.3% encountered issues related to excess or dysfunctional adiposity (stage 1) with a similar distribution between sexes. The proportion of the middle-aged population (45-64 years old) increased to 30.6% compared to this age group in stage 0. Over half of the US adults (56.5%) exhibited either metabolic risk factors, CKD, or a combination of both and met the criteria for inclusion in stage 2. 49.3% of the participants stage 2 were female and 50.7% were male. Older individuals were more prevalent in Stage 2: among the participants at stage 2 41.1% were between the ages of 45-64 and 20.4% aged 65 or older.

### Prevalence of CKM Syndrome Stages stratified by Sex, Age, and Race/Ethnicity

In comparison to females, males tended to experience CKM risk factors more frequently, with 19.2% of the overall male participants vs. 17.4% of the overall female participants exhibiting stage 1 CKM syndrome and 58.9% vs. 54.4% showing stage 2 (Figure 1A). Participants of advanced age were associated with higher CKM stage in US population without CVD. 81.1% of participants between 44 and 64 years and 69.6% of aged 65 years or older were attributed to either stage 1 or stage 2. While younger participants demonstrated a lower CKM risk, as illustrated in Figure 1B, a relatively high proportion of CKM stage 2 were still observed among those aged 18 to 24 years (35.4%) and those aged 25 to 44 years (49.9%). Relative to males, females at a younger age were associated with a lower CKM risk (age group 18-44 in stage 0-1: 4% of US adults vs. 2.4%), while females with increasing age showed an increasing proportion of CKM stage 2 with 18.8% of US adults aged 45 or older compared to 16% for males (Figure 1C, S3 Table). Across all race and ethnicity groups, the proportions of CKM Stage 0-2 were comparable (Figure 1D).

**Figure 1.**
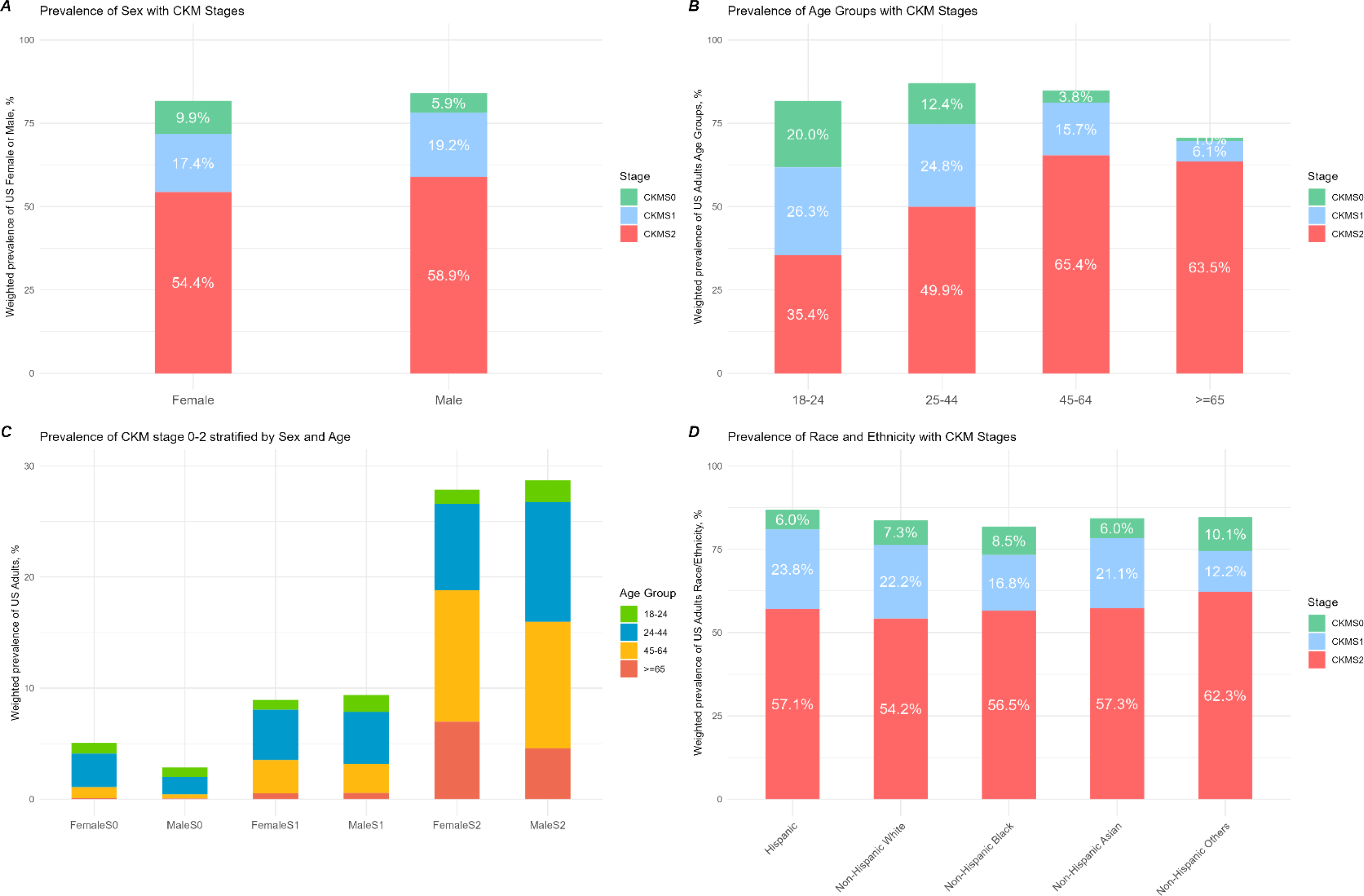
Sex-, Age– and race/ethnicity-stratified Prevalence in CKM Syndrome Stage 0-2. Sex-stratified prevalence of CKM syndrome stage 0-2 in US female adults or male adults (A), proportion of CKM syndrome stage 0-2 in different age groups (B), prevalence of CKM stage 0-2 in US adults stratified by combination of sex and age (C), proportion of CKM syndrome stage 0-2 in different race/ethnicity groups (D). All prevalence estimates are presented as weighted prevalence. Abbreviations: CKM, cardiovascular-kidney-metabolic syndrome.

### Prevalence of Metabolic Risk Factors and CKD in CKM Stage 2

Among all US adults with CKM stage 2, hypertension ranked as the most prevalent component (70.6%), followed by hypertriglyceridemia at 52.4% and metabolic syndrome (MetS) at 42.6%. The prevalence of CKD (16.4%) and diabetes (16.4%) were lower (Figure 2). S1-5 Figure illustrate the sex– and age-stratified prevalence of different components in CKM Stage 2. Overall, CKD was more common among females than males across all age groups, particularly in the age group of 65 years or older (23.6% of the participants at stage 2 with CKD vs. 13.9%, *P* < 0.001). At a younger age, females had a lower prevalence of hypertension compared to males (age group 18-24: 1.0% of the participants at stage 2 with hypertension vs. 2.4%, *P*<0.001; age group 25-44: 10.2% vs. 16.4%, *P* < 0.001). Hypertension was however more prevalent in women aged 65 or older (15.2% vs. 9.9%, *P* < 0.001). The prevalence distribution of hypertriglyceridemia, when stratified by sex and age, was similar to that of hypertension. Females aged 65 or older carried a higher burden of MetS (11.3% of the participants at stage 2 with MetS vs. 6.6%, *P* < 0.001). The prevalence of diabetes was similar among females and males in all age groups.

**Figure 2.**
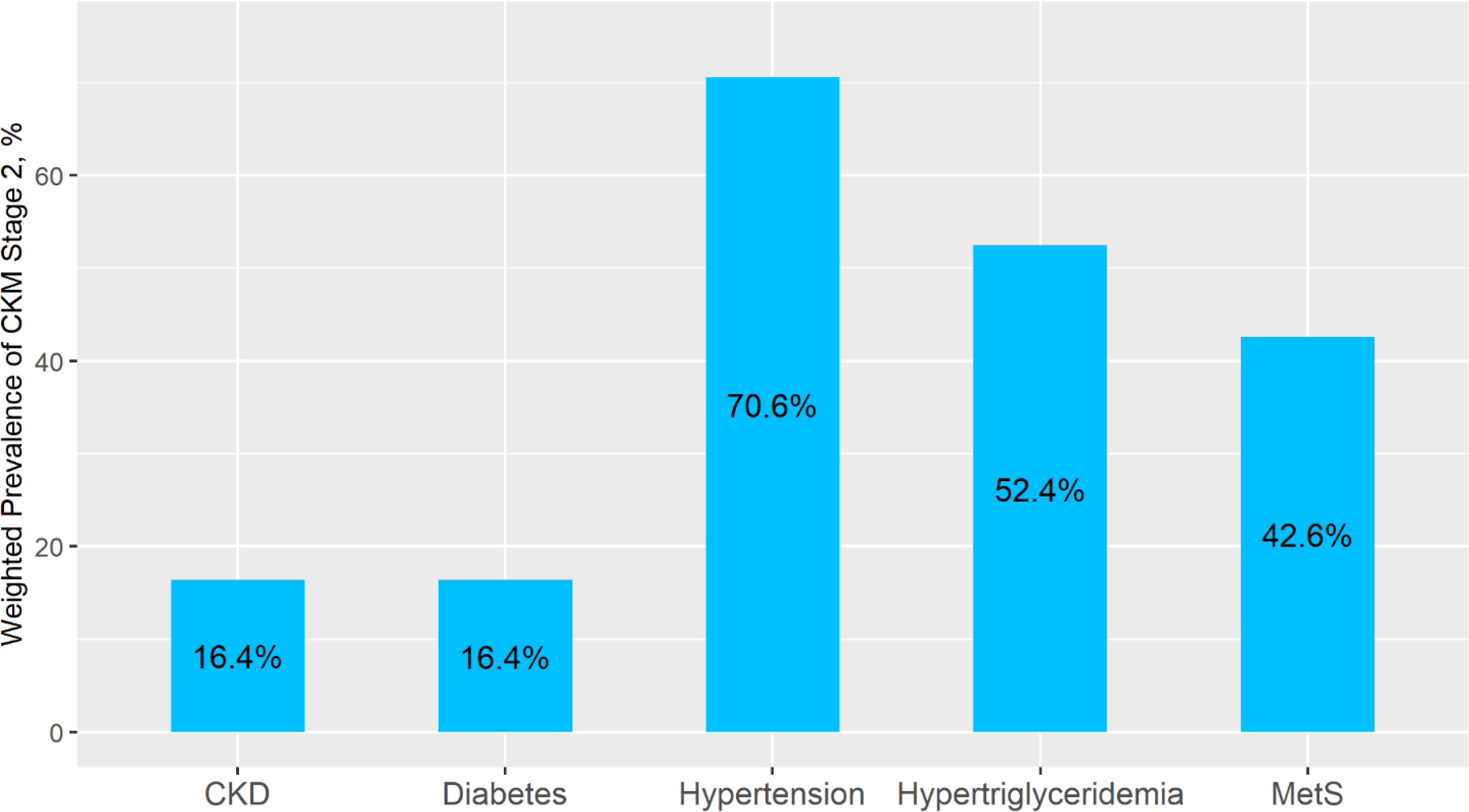
Overall Prevalence of CKD, Diabetes, Hypertension, Hypertriglyceridemia and MetS in US Adults with CKM syndrome Stage 2. Prevalence estimates are presented as weighted prevalence of noninstitutionalized US adults at stage 2. Abbreviations: CKM, cardiovascular-kidney-metabolic syndrome; CKD, chronic kidney disease; MetS, metabolic syndrome.

### Trends in Prevalence of CKM Stages

The comprehensive trend analysis for overall and sex-/age-stratified prevalence of CKM stages is summarized in Table 2. Overall, the proportion of US adult population in CKM stage 0 decreased from 7.9% in the 1999 to 2000 cycle to 6.8% in the 2017 to 2020 cycle (*P* for trend = 0.0018) (Figure 3). This declining trend was evident not only in females, with a reduction from 5.2% to 4.2% (*P* for trend= 0.0039), but also in males, with a slight decrease from 2.7% to 2.6% (*P* for trend = 0.031) (Figure 4). Additionally, there was a significant decrease in CKM stage 0 within the population aged 25 to 44 years (from 5.3% to 3.6%, *P* for trend < 0.001) (Figure 5A). For CKM stage 2, participants aged between 25 to 44 years demonstrated a substantial reduction from 23.2% to 16.8% (*P* for trend = 0.018), accompanied by a noteworthy increase in the proportion of participants aged 65 years or older (from 10.3% to 13.2%, *P* for trend < 0.001) (Figure 5C). The overall and sex-/age-stratified trends in CKM stage 1 remained relatively constant (Figure 3, Figure 4B and Figure 5B).

**Figure 3.**
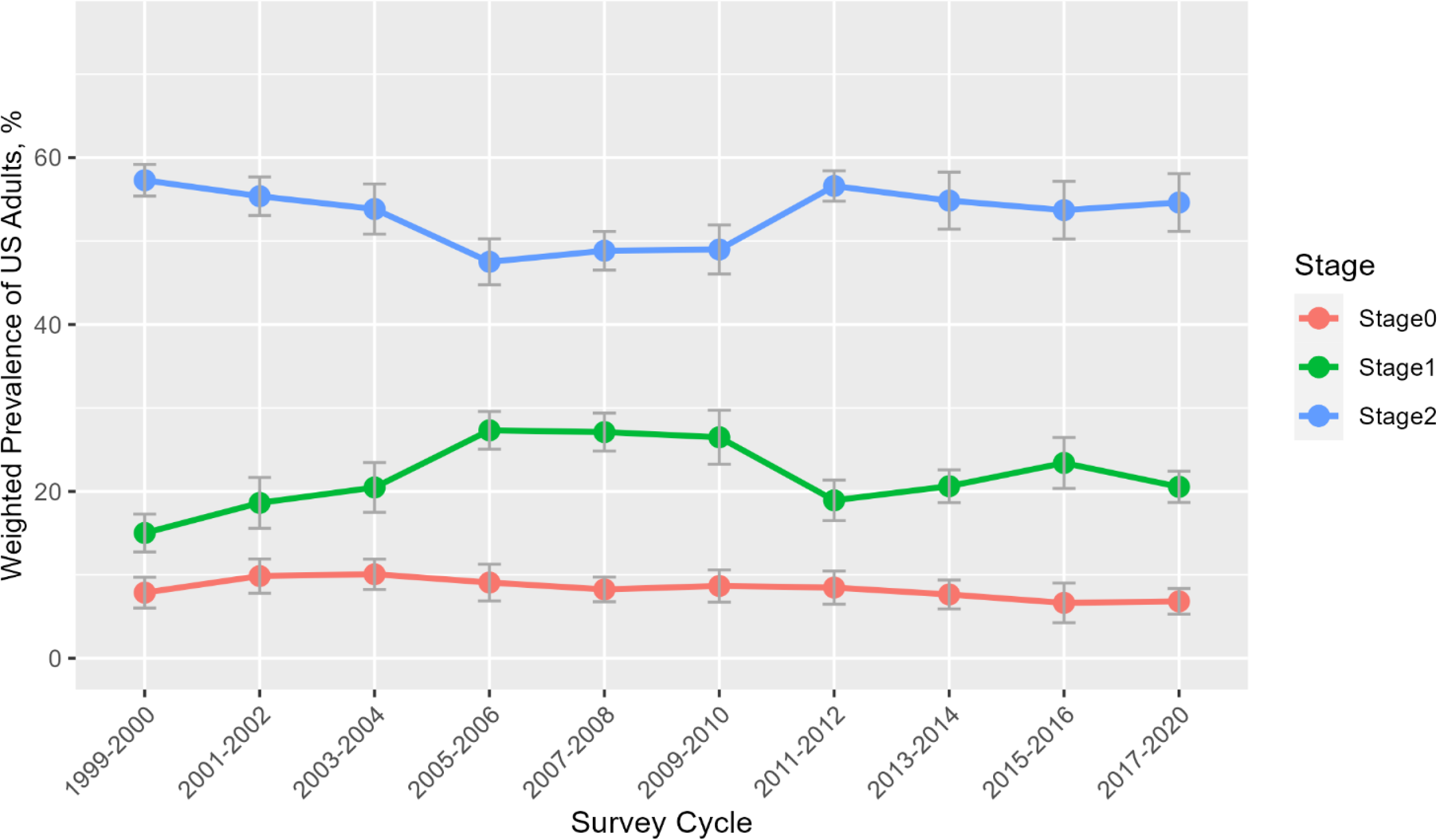
Trends in Prevalence of CKM Stage 0-2 among US adult Population, 1999-2000. CKM stage 0 *P* for trends = 0.0018; CKM stage 1 *P* for trends = 0.5; CKM stage 2 *P* for trends = 0.87. All prevalence estimates are presented as weighted prevalence of noninstitutionalized US adults. Error bars indicate 95% Cis CKM, cardiovascular kidney-metabolic syndrome.

**Figure 4.**
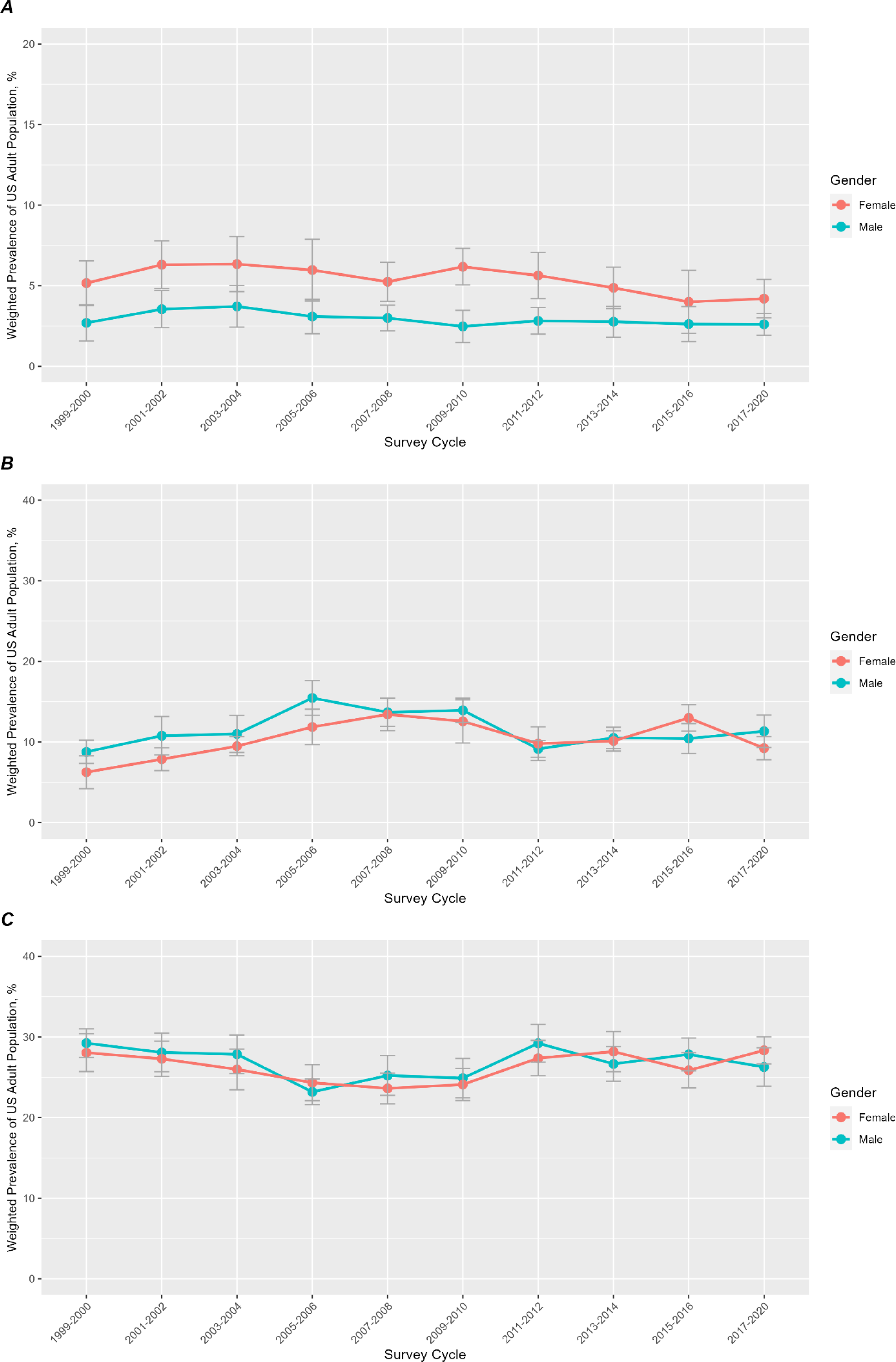
Trends in Sex-stratified Prevalence of CKM Stage 0-2, 1999-2020. CKM stage 0: female *P* for trends = 0.0039, male *P* for trends = 0.031; CKM stage 1: female *P* for trends = 0.18, male *P* for trends = 0.95; CKM stage 2: female *P* for trends = 0.77, male *P* for trends = 0.57. All prevalence estimates are presented as weighted prevalence of noninstitutionalized US adults. Error bars indicate 95% CIs. CKM, cardiovascular kidney-metabolic syndrome.

**Figure 5.**
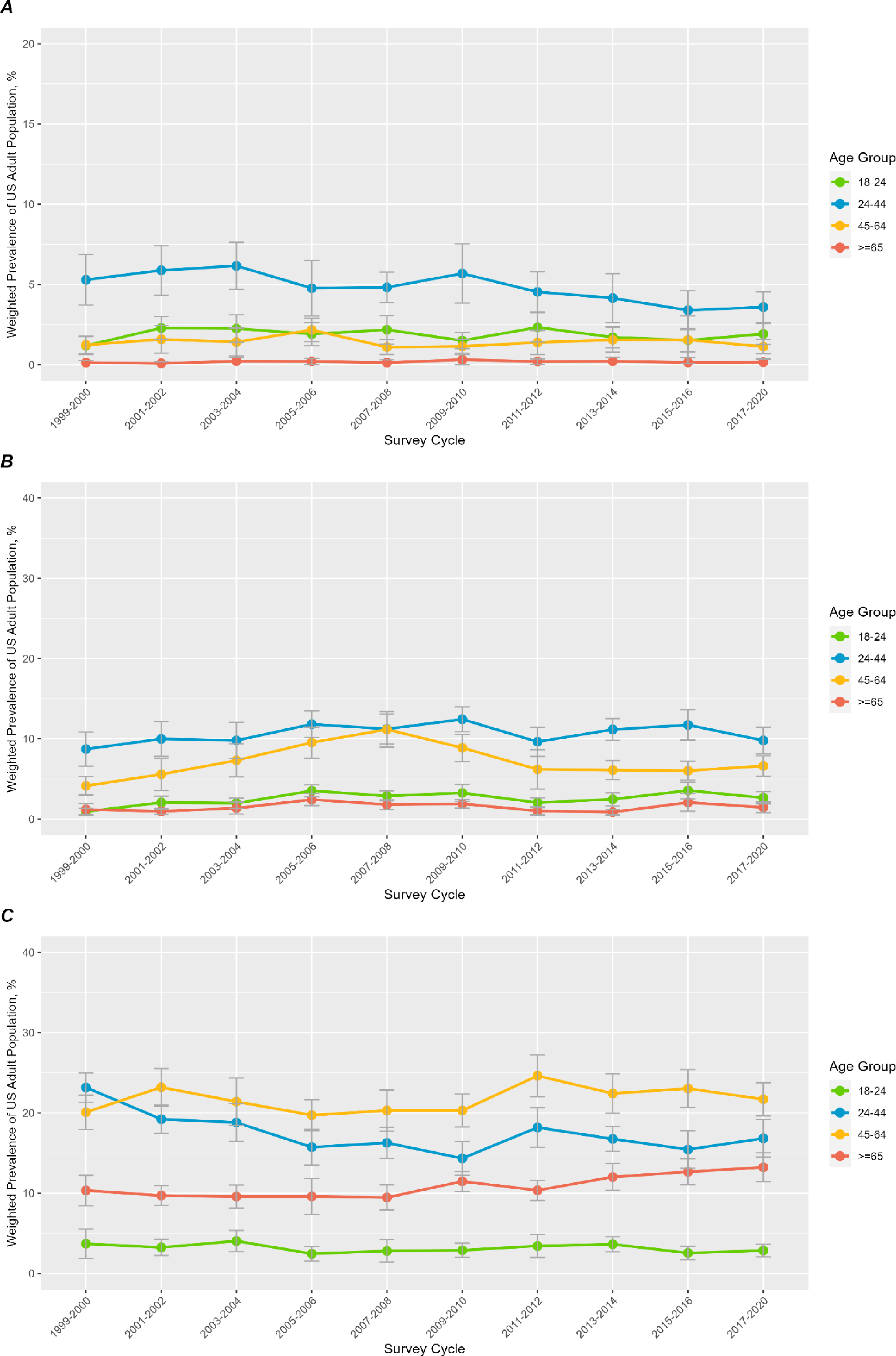
Trends in Age-stratified Prevalence of CKM Stage 0-2, 1999-2020. CKM stage 0: age 18-24 *P* for trends = 0.88, age 24-44 *P* for trends < 0.001, age 45-64 *P* for trends = 0.58, age≥65 *P* for trends = 0.53; CKM stage 1: age 18-24 *P* for trends = 0.079, age 24-44 *P* for trends = 0.36, age 45-64 *P* for trends = 0.89, age≥65 *P* for trends = 0.75; CKM stage 2: age 18-24 *P* for trends = 0.2, age 24-44 *P* for trends = 0.018, age 45-64 *P* for trends = 0.33, age≥65 *P* for trends < 0.001. All prevalence estimates are presented as weighted prevalence of noninstitutionalized US adults. Error bars indicate 95% CIs. CKM, cardiovascular kidney-metabolic syndrome.

**Table 2.** Trends in weighted Overall and Sex-/Age-stratified Prevalence of CKM stages in US Adults, 1999-2020. Abbreviations: CKM, cardiovascular-kidney-metabolic syndrome. *Estimate may be unreliable with relative standard error >30%

|  | Weighted Prevalence, % (95% CI) |  |  |  |  |  |  |  |  |  | P value |
| --- | --- | --- | --- | --- | --- | --- | --- | --- | --- | --- | --- |
| Characteristic | 1999-2000<br>n= 186356015 | 2001-2002<br>n= 195531851 | 2003-2004<br>n=201768320 | 2005-2006<br>n= 204518688 | 2007-2008<br>n= 212681297 | 2009-2010<br>n= 216303553 | 2011-2012<br>n= 221814830 | 2013-2014<br>n= 227000000 | 2015-2016<br>n= 231226225 | 2017-2020<br>n= 237963984 |  |
| CKM Stage 0 | 7.9 (6.0 -9.7) | 9.9 (7.8 - 11.9) | 10.1 (8.2 - 11.9) | 9.1 (6.9 - 11.3) | 8.2 (6.8 - 9.7) | 8.7 (6.7 - 10.60) | 8.5 (6.5 - 10.4) | 7.6 (5.9 - 9.4) | 6.6 (4.3 - 9.0) | 6.8 (5.3 - 8.3) | 0.0018 |
| CKM Stage 1 | 15.0 (12.8 - 17.3) | 18.6 (15.6 - 21.7) | 20.5 (17.5 - 23.5) | 27.3 (25.1 - 29.6) | 27.1 (24.8 - 29.4) | 26.5 (23.3 - 29.7) | 18.9 (16.5 - 21.4) | 20.6 (18.7 - 22.6) | 23.4 (20.4 - 26.5) | 20.6 (18.7 - 22.4) | 0.5 |
| CKM Stage 2 | 57.3 (55.4-59.2) | 55.4 (53.1 - 57.5) | 53.8 (50.8 - 56.8) | 47.5 (44.8 - 50.3) | 48.8 (46.5 - 51.2) | 49.0 (46.1 - 51.9) | 56.6 (54.8 - 58.40) | 54.8 (51.4 - 58.3) | 53.7 (50.2 - 57.2) | 54.6 (51.2 - 58.1) | 0.87 |
| **CKM Stage 0** |  |  |  |  |  |  |  |  |  |  |  |
| Female | 5.2 (3.8 - 6.5) | 6.3 (4.8 - 7.8) | 6.3 (4.6 - 8.1) | 6.0 (4.1 - 7.9) | 5.2 (4.0 - 6.5) | 6.2 (5.0 - 7.3) | 5.6 (4.3 - 7.1) | 4.9 (3.6 - 6.2) | 4.0 (2.1 - 6.0) | 4.2 (3.0 - 5.4) | 0.0039 |
| Male | 2.7 (1.6 - 3.8) | 3.5 (2.4 - 4.7) | 3.7 (2.4 - 5.0) | 3.1 (2.0 - 4.3) | 3.0 (2.2 - 3.8) | 2.5 (1.5 - 3.5) | 2.8 (2.0 - 3.7) | 2.8 (1.8 - 3.7) | 2.6 (1.5 - 3.7) | 2.6 (1.9 - 3.3) | 0.031 |
| Age 18 - 24 | 1.2 (0.6 - 1.8) | 2.3 (1.6 - 3.0) | 2.3 (1.4 - 3.1) | 1.9 (1.2 - 2.6) | 2.2 (1.3 - 3.1) | 1.5 (1.0 - 2.0) | 2.3 (1.4 - 3.2) | 1.7 (1.1 - 2.4) | 1.5 (0.8 - 2.2) | 1.9 (1.3 - 2.6) | 0.88 |
| Age 25 - 44 | 5.3 (3.7 - 6.9) | 5.9 (4.3 - 7.4) | 6.2 (4.7 - 7.6) | 4.8 (3.0 - 6.5) | 4.8 (3.9 - 5.8) | 5.7 (3.8 - 7.5) | 4.5 (3.3 - 5.8) | 4.2 (2.6 - 5.7) | 3.4 (2.2 - 4.6) | 3.6 (2.6 - 4.5) | < 0.001 |
| Age 45 - 64 | 1.2 (0.7 - 1.8) | 1.6 (0.7 - 2.4) | 1.4 (0.6 - 2.3) | 2.2 (1.4 - 2.9) | 1.1 (0.6 - 1.6) | 1.1 (0.7 - 1.6) | 1.4 (0.6 - 2.1) | 1.6 (0.8 - 2.3) | 1.6 (0.1 - 3.0) | 1.1 (0.7 - 1.6) | 0.58 |
| *Age >= 65 | 0.1 (0.0 - 0.3) | 0.1 (0.0 - 0.2) | 0.2 (0.0 - 0.5) | 0.2 (0.0 - 0.4) | 0.1 (0.0 - 0.3) | 0.3 (0.0 - 0.6) | 0.2 (0.0 - 0.4) | 0.2 (0.0 - 0.5) | 0.1 (0.0 -0.4) | 0.2 (0.0 - 0.4) | 0.53 |
| **CKM Stage 1** |  |  |  |  |  |  |  |  |  |  |  |
| Female | 6.2 (4.2 - 8.3) | 7.9 (6.4 - 9.3) | 9.5 (8.3 - 10.7) | 11.9 (9.7 - 14.1) | 13.4 (11.4 - 15.4) | 12.56 (9.9 - 15.2) | 9.8 (7.7 - 11.9) | 10.1 (8.9 - 11.4) | 13.0 (11.3 - 14.6) | 9.2 (7.8 - 10.7) | 0.18 |
| Male | 8.8 (7.3 - 10.2) | 10.8 (8.4 - 13.2) | 11.0 (8.7 - 13.3) | 15.5 (13.3 - 17.6) | 13.7 (11.9 - 15.4) | 13.9 (12.4 - 15.4) | 9.1 (8.1 - 10.2) | 10.5 (9.2 - 11.8) | 10.4 (8.6 - 12.3) | 11.3 (9.3 - 13.3) | 0.95 |
| Age 18 - 24 | 0.9 (0.5 - 1.4) | 2.1 (1.3 - 2.9) | 2.0 (1.3 - 2.6) | 3.5 (2.8 - 4.3) | 2.9 (2.3 - 3.5) | 3.3 (2.2 - 4.2) | 2.1 (1.5 - 2.7) | 2.5 (1.6 - 3.3) | 3.6 (2.5 - 4.6) | 2.7 (1.9 - 3.4) | 0.079 |
| Age 25 - 44 | 8.7 (6.6 - 10.9) | 10.0 (7.8 - 12.2) | 9.8 (7.5 - 12.1) | 11.8 (10.2 - 13.5) | 11.2 (9.4 - 13.1) | 12.4 (10.9 - 14.0) | 9.6 (7.8 - 11.5) | 11.2 (9.8 - 12.5) | 11.7 (9.8 - 13.6) | 9.8 (8.1 - 11.5) | 0.36 |
| Age 45 - 64 | 4.1 (3.0 - 5.3) | 5.6 (3.6 - 7.6) | 7.3 (5.2 - 9.4) | 9.5 (7.6 - 11.5) | 11.2 (8.9 - 13.4) | 8.9 (7.2 - 10.6) | 6.2 (3.8 - 8.6) | 6.1 (4.9 - 7.3) | 6.0 (4.9 - 7.2) | 6.6 (5.4 - 7.9) | 0.89 |
| Age >= 65 | 1.2 (0.5 - 2.0) | 1.0 (0.6 - 1.3) | 1.4 (0.6 - 2.1) | 2.4 (1.7 - 3.2) | 1.8 (1.2 - 2.4) | 1.9 (1.4 - 2.4) | 1.0 (0.5 - 1.5) | 0.9 (0.5 - 1.2) | 2.1 (1.0 - 3.2) | 1.5 (0.8 - 2.1) | 0.75 |
| **CKM Stage 2** |  |  |  |  |  |  |  |  |  |  |  |
| Female | 28.1 (25.7 - 30.4) | 27.3 (25.1 - 29.5) | 26.0 (23.5 - 28.5) | 24.3 (22.1 - 26.6) | 23.6 (21.7 - 25.5) | 24.1 (22.1 - 26.1) | 27.4 (25.2 - 29.6) | 28.2 (25.7 - 30.7) | 25.9 (23.7 - 28.1) | 28.3 (26.7 - 30.0) | 0.77 |
| Male | 29.2 (27.5 - 31.0) | 28.1 (25.7 - 30.5) | 27.9 (25.5 - 30.3) | 23.2 (21.6 - 24.8) | 25.2 (22.8 - 27.7) | 24.9 (22.4 - 27.3) | 29.2 (26.9 - 31.6) | 26.7 (24.5 - 28.8) | 27.8 (25.8 - 29.9) | 26.3 (23.9 - 28.7) | 0.57 |
| Age 18 - 24 | 3.7 (1.9 - 5.5) | 3.2 (2.2 - 4.3) | 4.0 (2.7 - 5.4) | 2.5 (1.5 - 3.4) | 2.8 (1.4 - 4.2) | 2.9 (2.0 - 3.8) | 3.4 (2.0 - 4.9) | 3.6 (2.7 - 4.6) | 2.6 (1.7 - 3.4) | 2.9 (2.1 - 3.7) | 0.2 |
| Age 25 - 44 | 23.2 (21.4 - 25.0) | 19.2 (17.5 - 21.0) | 18.8 (16.4 - 21.2) | 15.7 (13.5 - 18.0) | 16.3 (14.3 - 18.2) | 14.3 (12.2 - 16.4) | 18.2 (15.7 - 20.7) | 16.8 (15.2 - 18.3) | 15.4 (13.1 - 17.8) | 16.8 (14.5 - 19.2) | 0.018 |
| Age 45 - 64 | 20.1 (17.9 - 22.2) | 23.2 (20.9 - 25.5) | 21.4 (18.5 - 24.4) | 19.7 (17.8 - 21.7) | 20.3 (17.7 - 22.9) | 20.3 (18.2 - 22.4) | 24.6 (22.0 - 27.2) | 22.4 (20.0 - 24.9) | 23.0 (20.7 - 25.4) | 21.7 (19.6 - 23.8) | 0.33 |
| Age >= 65 | 10.3 (8.4-12.2) | 9.7 (8.5 - 10.9) | 9.6 (8.2 - 11.0) | 9.6 (7.3 - 11.8) | 9.5 (7.9 - 11.0) | 11.5 (10.2 - 12.7) | 10.3 (9.1 - 11.6) | 12.0(10.3-13.7) | 12.7 (11.0 - 14.3) | 13.2 (11.4 - 15.0) | < 0.001 |

### Population with undefined CKM Stages

During the selection process for each CKM stage, we identified specific subgroups that remained undefined. This included participants, for example, only with abnormal HDL-cholesterol or LDL-cholesterol. These participants fulfill neither the definition of stage 0 (normal lipid profile) nor of stage 1. As depicted in Figure 6, participants with undefined stage comprised 3.3% of the overall US adult population, with 62.3% (4.0% of the overall female adult population) being female and 37.7% (2.6% of the overall male adult population) being male. After incorporating this proportion of undefined subgroup into CKM stages 0-2, the sex-stratified prevalence of CKM stages 0-2 became comparable.

**Figure 6.**
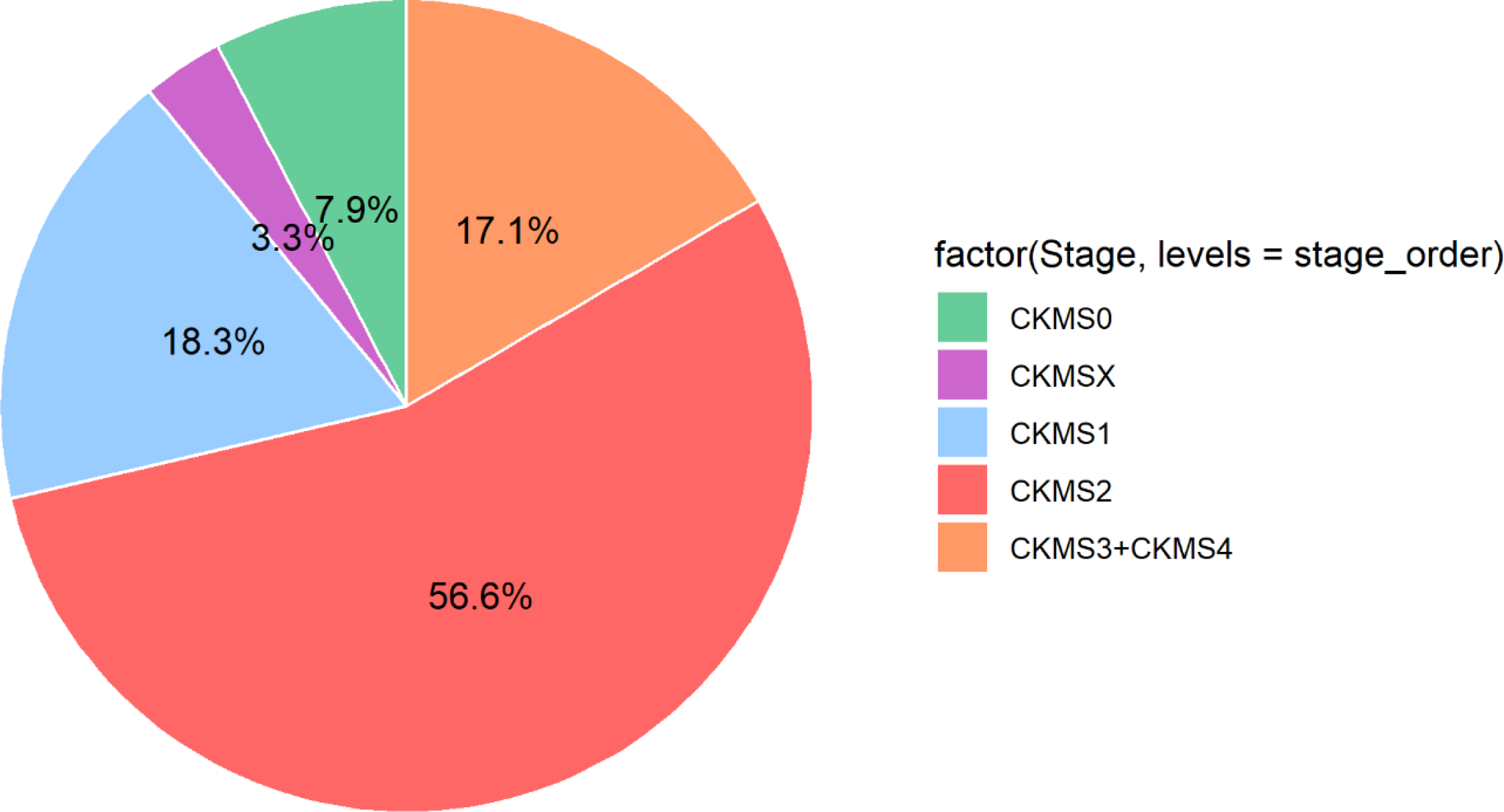
Prevalence of CKM Syndrome Stage 0-1 and undefined stage (CKM SX) in US Adults. Prevalence estimates are shown as weighted prevalence of noninstitutionalized US adults. CKMSX represents US adults, who did not yet fulfill the criterion for CKM stage 1 but had abnormalities excluding them from stage 0. Abbreviations: CKM, cardiovascular-kidney-metabolic syndrome; S0, stage 0; SX, undefined stage; S1, stage 1; S2, stage 2.

### Sensitivity Analysis

After reweighting for nonresponse, we observed no significant deviation from our main analysis (S4 Table). The aggregated outcome from 5 imputations, following the imputation of missing data, aligned consistently with the main analysis. (S5 Table).

## Discussion

Our analysis of this nationally representative survey of US adults reveals a high prevalence of CKM syndrome in the general population. Signs of CKM syndrome were absent in only less than 10% of US adults. Even among younger adults, we observed a higher burden of CKM syndrome: 1 in 4 US adults aged between 18-24 years had stage 1, and 1 in 3 exhibited stage 2. Women aged 65 or older were at the highest risk for CKM syndrome stage 2. From 1999-2000 through 2017-2020, the prevalence of CKM stage 0 decrease from 7.9% to 6.8%. Furthermore, the prevalence of various metabolic risk factors and CKD in CKM syndrome stage 2 varied across sex and age.

A prior study, which examined cardiovascular disease, CKD, and diabetes among US adults, using also the NHANES survey, showed a high and increasing prevalence of cardiac, renal and metabolic multimorbidity ^13^. Our analysis further confirms the high burden of CKM syndrome after applying defined criteria for each stage. However, the focus is on the population at stages 0-2, who have not yet developed subclinical or clinical CVD, to identify the demographic subsets who would profit from interdisciplinary comprehensive preventive strategies for achieving a reduction in cardiovascular and mortality risks.

CKM syndrome is an evolving condition that begins already in childhood ^29,30^. Current evidence demonstrates that trends in CKM syndrome have been increasing in young adulthood despite the general improvement of health care^1,31–33^. In this analysis, we observed a high prevalence of poor CKM health in younger US adult population, with CKM syndrome absent in only 20% of adults aged between 18-24 years and 12.4% adults aged between 25-44 years. Over the past two decades, younger adults have exhibited a significant decreasing trend in the prevalence of stage 0, demonstrating a shift towards less favorable CKM health. These observations imply an escalating burden of CKM syndrome in the future as these younger demographic ages, unless the adverse trends in the prevalence of CKM risk factors can be reversed. To reinforce initiatives promoting the CKM health of young adults, the public health policymakers should comprehend the unique interplay of biological, interpersonal, socioeconomic disparities and behavioral features that characterize this life stage. For instance, young adults could be more motivated to access their health records using mobile/wearable devices. Healthcare providers could, therefore, collaborate with patients to provide personalized medical advice by integrating these digital data ^34,35^. In addition, more CKM health trials should be conducted with a focus on including participants under 45 years old, as young adults have been underrepresented in behavioral and pharmacologic prevention/ intervention trials so far ^36–39^.

For the effective management of CKM syndrome, it’s essential to also consider sex and age disparities in order to prevent a progression into stage 3-4. In CKM syndrome stage 2, for example, as women reached an older age (≥ 65 years), they showed a significant higher prevalence across almost all metabolic risk factors and CKD compared to men in the same age group. Notably, this higher prevalence persisted in CKD, regardless of age. According to US Renal Data system 2023 annual Report, women have experienced more often CKD than men since 2005-2008 ^12^. Several studies demonstrated a significant deterioration in lipid profiles and an increasing prevalence of MetS with menopause, beyond the effects of chronological aging, leading to a remarkable CKM progression and thus an increase of CVD absolute risk after menopause ^40–43^. We also observed that men at younger age had significant higher prevalence of hypertension, MetS and hypertriglyceridemia. To address this unmet need referred to sex– and age-disparities, additional efforts should be directed towards research fields, clinical practice, and guideline development.

Despite the detailed definition of each CKM syndrome stage, a specific population remained undefined, who had one or two abnormal lipid parameters. For an optimal risk assessment, accurately categorizing this subgroup population is essential. Besides, current evidence demonstrated a U-shaped correlation between HDL-cholesterol and cardiovascular risk ^44–47^. For classification of CKM syndrome stages, very high HDL-cholesterol level should be considered as a risk factor. Furthermore, CKM syndrome stage 2 encompasses a broad spectrum of chronic conditions and includes more than half of the US adult population. Following the implementation of an enhanced screening system, a certain segment population from stage 2 may be categorized into stage 3. Concurrently, a refined risk assessment tool should be integrated with the CKM staging system to identify individuals at higher risk of CVD, with the goal of ensuring individualized healthcare. The new PREVENT model incorporates not only the traditional CVD risk factors but also kidney function along with other predictors such as UACR, HbA1c, and social determinants, as additional add-on model approach ^9^. We propose defining a new CKM syndrome stage for individuals in stage 2 but at higher risk of CVD after applying PREVENT risk assessment, positioned between stage 2 and stage 3.

This study has several limitations. First, it comprises only the noninstitutionalized, civilian population, thus does not capture the individuals in nursing homes or the military. Second, non-Hispanic Asian participants were not classified and oversampled until 2011. This subgroup population might be under-represented in our study. Third, we didn’t distinguish between diabetes mellitus types. This decision was based on our opinion that type 1 diabetes should also be included in CKM syndrome due to its association with CKD progression and higher cardiovascular risk. Finally, nonresponse might substantially influence our conclusion. However, sensitivity analyses, one after reweighting for nonresponse and another after imputing missing data, demonstrated comparable results, which confirmed our main analysis.

## Conclusion

The definition of CKM syndrome is an important step in increasing awareness for the deleterious interplay of obesity, metabolic and renal factors in causing cardiovascular sequelae. Our study illustrates an exceptionally high burden of CKM syndrome among US adults, mirroring trends observed in all Western countries. Given the high prevalence of stage 2 of CKM syndrome, additional risk assessment is mandatory to tailor preventive measures on the population level to efficiently reduce cardiovascular morbidity and mortality.

## Data Availability

NHANES data are available from the NHANES web page. The R code of the analysis is available from the authors on reasonable request.

## Abbreviations and Acronyms

BMI: Body mass index
CKD: Chronic kidney disease
CKM: syndrome Cardiovascular-kidney-metabolic syndrome
CVD: Cardiovascular disease
eGFR: Estimated glomerular filtration rate
MetS: Metabolic syndrome
NHANES: National Health and Nutrition Examination Survey
UACR: Urinary albumin to creatinine ratio

## Acknowledgements

No other individuals contributed to this study substantively.

